# Reduced control of SARS-CoV-2 infection is associated with lower mucosal antibody responses in pregnant women

**DOI:** 10.1101/2023.03.19.23287456

**Authors:** Laura A. St Clair, Raghda E. Eldesouki, Jaiprasath Sachithanandham, Anna Yin, Amary Fall, C. Paul Morris, Julie M. Norton, Michael Forman, Omar Abdullah, Santosh Dhakal, Caelan Barranta, Hana Golding, Susan J. Bersoff-Matcha, Catherine Pilgrim-Grayson, Leah Berhane, Andrea L. Cox, Irina Burd, Andrew Pekosz, Heba H. Mostafa, Eili Y. Klein, Sabra L. Klein

**Author notes:** **Corresponding authors:**, 5801 Smith Ave, Davis Suite 3220, Baltimore, MD 21209, **(Primary)**, 615 North Wolfe Street, Rm W2118, Baltimore, MD 21205. equal contributors (Laura A. St Clair, Raghda E. Eldesouki, Jaiprasath Sachithanandham, Anna Yin). co-senior authors (Eili Y. Klein, Sabra L. Klein).

## Abstract

**Importance:** Pregnant women are at increased risk of severe COVID-19, but the contribution of viral RNA load, the presence of infectious virus, and mucosal antibody responses remain understudied.

**Objective:** To evaluate the association of COVID-19 outcomes following confirmed infection with vaccination status, mucosal antibody responses, infectious virus recovery and viral RNA levels in pregnant compared with non-pregnant women.

**Design:** A retrospective observational cohort study of remnant clinical specimens from SARS-CoV-2 infected patients between October 2020-May 2022.

**Setting:** Five acute care hospitals within the Johns Hopkins Health System (JHHS) in the Baltimore, MD-Washington, DC area.

**Participants:** Participants included confirmed SARS-CoV-2 infected pregnant women and matched non-pregnant women (matching criteria included age, race/ethnicity, and vaccination status).

**Exposure:** SARS-CoV-2 infection, with documentation of SARS-CoV-2 mRNA vaccination.

**Main Outcome(s):** The primary dependent measures were clinical COVID-19 outcomes, infectious virus recovery, viral RNA levels, and mucosal anti-spike (S) IgG titers from upper respiratory tract samples. Clinical outcomes were compared using odds ratios (OR), and measures of virus and antibody were compared using either Fisher’s exact test, two-way ANOVA, or regression analyses. Results were stratified according to pregnancy, vaccination status, maternal age, trimester of pregnancy, and infecting SARS-CoV-2 variant.

**Results(s):** A total of 452 individuals (117 pregnant and 335 non-pregnant) were included in the study, with both vaccinated and unvaccinated individuals represented. Pregnant women were at increased risk of hospitalization (OR = 4.2; CI = 2.0-8.6), ICU admittance, (OR = 4.5; CI = 1.2-14.2), and of being placed on supplemental oxygen therapy (OR = 3.1; CI =1.3-6.9). An age-associated decrease in anti-S IgG titer and corresponding increase in viral RNA levels (*P*< 0.001) was observed in vaccinated pregnant, but not non-pregnant, women. Individuals in their 3^rd^ trimester had higher anti-S IgG titers and lower viral RNA levels (*P*< 0.05) than those in their 1^st^ or 2^nd^ trimesters. Pregnant individuals experiencing breakthrough infections due to the omicron variant had reduced anti-S IgG compared to non-pregnant women (*P*< 0.05).

**Conclusions and Relevance:** In this cohort study, vaccination status, maternal age, trimester of pregnancy, and infecting SARS-CoV-2 variant were each identified as drivers of differences in mucosal anti-S IgG responses in pregnant compared with non-pregnant women. Observed increased severity of COVID-19 and reduced mucosal antibody responses particularly among pregnant participants infected with the Omicron variant suggest that maintaining high levels of SARS-CoV-2 immunity may be important for protection of this at-risk population.

**Key Points:** *Question:* Is greater COVID-19 disease severity during pregnancy associated with either reduced mucosal antibody responses to SARS-CoV-2 or increased viral RNA levels?

*Finding:* In a retrospective cohort of pregnant and non-pregnant women with confirmed SARS-CoV-2 infection, we observed that (1) disease severity, including ICU admission, was greater among pregnant than non-pregnant women; (2) vaccination was associated with reduced recovery of infectious virus in non-pregnant women but not in pregnant women; (3) increased nasopharyngeal viral RNA levels were associated with reduced mucosal IgG antibody responses in pregnant women; and (4) greater maternal age was associated with reduced mucosal IgG responses and increased viral RNA levels, especially among women infected with the Omicron variant.

*Meaning:* The findings of this study provide novel evidence that, during pregnancy, lower mucosal antibody responses are associated with reduced control of SARS-CoV-2, including variants of concern, and greater disease severity, especially with increasing maternal age. Reduced mucosal antibody responses among vaccinated pregnant women highlight the need for bivalent booster doses during pregnancy.

## Introduction

The ongoing COVID-19 pandemic has caused more than 650 million confirmed SARS-CoV-2 cases and greater than 6.6 million deaths reported worldwide [1]. Pregnant women are classified as an at-risk group for severe complications, but the relationship between physiologic, immunologic, and hormonal changes that occur during pregnancy and increased disease severity risk remain unclear [2-4]. Analyses from the US Centers for Disease Control and Prevention (CDC), show that among people with confirmed SARS-CoV-2 infections from January 2020-December 2021, pregnant women were 5 times more likely to be admitted to an intensive care unit (ICU), had a 76% greater risk of requiring invasive ventilation, and had a 3.3 times greater risk of death compared to non-pregnant women [5]. Despite these increased risks, the immune responses to SARS-CoV-2 infection and efficacy of SARS-CoV-2 vaccination in pregnant women remains understudied [6-10]. Studies that have analyzed immune responses to SARS-CoV-2 infection and vaccination have largely focused on serological immunity, with limited analysis of the mucosal antibody response to SARS-CoV-2 infection [11] and its association with virus load, especially among pregnant women.

In this retrospective observational cohort study, remnant nasopharyngeal (NP) swab or lateral mid-turbinate nasal swab samples from pregnant and matched non-pregnant patients with confirmed positive SARS-CoV-2 infection who visited the Johns Hopkins Health System between October 2020-May 2022 were analyzed for clinical outcomes, virus lineage, infectious virus recovery, quantification of viral RNA level, and assessment of mucosal anti-spike (S) IgG titers. Differences in each measure were compared between non-pregnant and pregnant women and stratified by vaccination status, age, trimester of pregnancy, and infecting SARS-CoV-2 variants.

## Materials and Methods

### Ethical considerations and data availability

This study was conducted under the Johns Hopkins University IRB protocols IRB00221396, IRB00288258, IRB00289116 and a waiver of consent. Remnant clinical specimens from individuals who tested positive for SARS-CoV-2 following standard of care or diagnostic screening were used in this study. Whole viral genome sequencing was performed for genomic SARS-CoV-2 surveillance and sequences were made publicly available in the GISAID database.

### Subjects and sample selection

This was a retrospective observational cohort study that used remnant nasopharyngeal swabs (from symptomatic patients) or lateral mid-turbinate nasal swabs (from asymptomatic patients) after standard of care diagnostic screening for SARS-CoV-2 infection across JHHS. Multiple molecular assays were performed to confirm SARS-CoV-2 infection, as previously described [12, 13]. Clinical information, and information about vaccination and immune status of individuals were bulk extracted from the electronic health record that is shared across JHHS and analyzed as previously described in [14]. The cohort excluded anyone who identified as male, whose sex at birth was recorded as male, or who chose not to disclose their sex at birth. Propensity score matching was used to select a cohort of control patients (3:1 ratio of control to pregnant patients). Psmatch2 in Stata was used to match the patients on the variables listed above using two methods, the first used no replacement (i.e., selection of best matches for every pregnant patient in the cohort), then with a nearest neighbor of 4 with a caliper of 0.01 was used to select additional patients that might be near close matches. Initial selection identified 287 pregnant patients and 817 matched non-pregnant controls; however, of this group, complete vaccination data, full sequencing data, and remnant clinical specimens were only available for 117 pregnant individuals (84 unvaccinated, 33 vaccinated), and 335 matched non-pregnant controls (244 unvaccinated, 91 vaccinated) which defined the final cohort (**Table 1**). For the purposes of this study, vaccinated individuals were defined as those who either received two primary doses (Pfizer/BioNTech or Moderna mRNA-1273 vaccines) or received the primary doses and third booster dose prior to confirmed infection. Unvaccinated individuals were defined as individuals who had received no COVID-19 vaccine prior to infection.

**Table 1.**
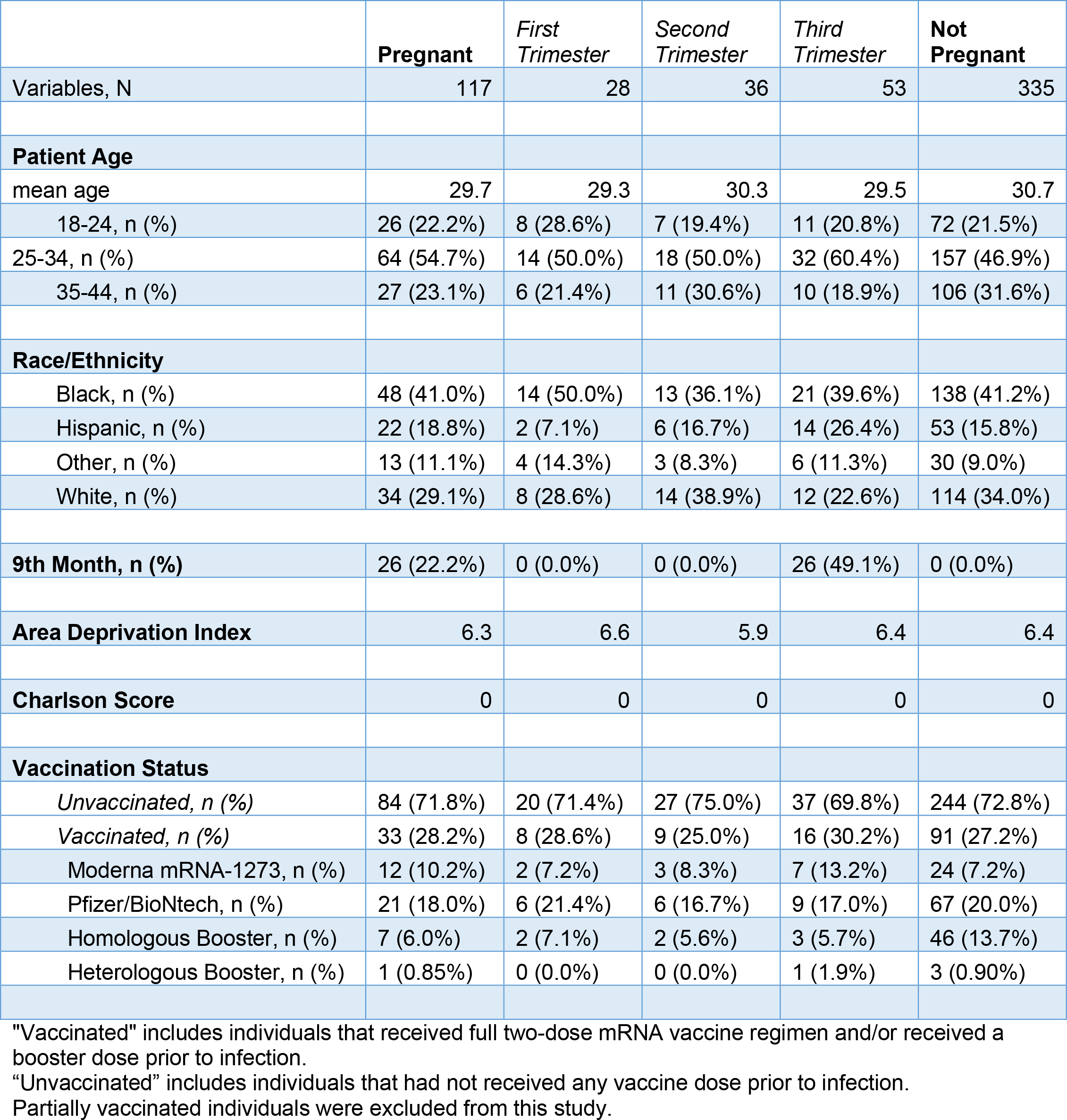
Patients and samples used in this study.

Individuals who were partially vaccinated were excluded from this study.

### Amplicon-based Sequencing

Specimen preparation, extractions, and sequencing were performed as described previously [15, 16]. NEBNext® ARTIC SARS-CoV-2 Companion Kit (VarSkip Short SARS-CoV-2 # E7660-L) was used for library preparation and sequencing using the Nanopore GridION. Base-calling of reads was conducted using the MinKNOW, followed by demultiplexing with guppybarcoder that requires barcodes at both ends. Artic-ncov2019 medaka protocol was used for alignment and variant calling [17, 18]. Clades were determined using Nextclade beta v 1.13.2 (clades.nextstrain.org, Last accessed March 30, 2022), and lineages were determined with Pangolin COVID-19 lineage Assigner [19]. Sequences with coverage >90% and mean depth >100 were submitted to GISAID database.

### SARS-CoV-2 PCR

After clinical diagnosis, samples were retested using the CDC designed primers and probes for the N gene to assess viral RNA levels (Cycle threshold, or Ct) [20]. Equivalent distribution of data between samples collected from NP swabs and lateral mid-turbinate nasal swabs was observed; as such, analysis of Ct values did not control for sample type.

### Infectious SARS-CoV-2 recovery

TMPRSS2 VeroE6 cells (RRID: CVCL_YQ49) obtained from the cell repository of the National Institute of Infectious Diseases, Japan [18, 21], and were cultured as previously described [22]. For virus isolation, cells plated in 24-well dishes had the culture media replaced with 350 µL of infection media (culture media except that the FBS was reduced to 2.5%), followed by the addition of 150 µL of swab specimen. After incubation for 2 hours at 37°C, the inoculum was removed and replaced with 500 µL infection media. The cells were monitored daily for the appearance of SARS-CoV-2 cytopathic effect (CPE) and the presence of SARS-CoV-2 genomes in CPE positive samples was confirmed by reverse transcriptase PCR (RT-PCR) as previously described [23].

### Indirect enzyme-linked immunosorbent assays

The protocol was adapted from published protocols [24] that were established previously in our laboratory [6, 25-27], and was modified to assess total IgG from viral transport media (VTM). Briefly, 96-well plates were coated with full-length vaccine-strain (ancestral) Spike (S) protein (SeroNet) and incubated overnight at 4°C. Coating buffer was removed, plates were washed, and blocked for 1 hour at room temperature. All samples were heat inactivated at 56°C for 1 hour prior to use. Negative controls using pooled VTM from COVID-19 negative patients were plated at final concentration of 1:4. Positive control samples using a monoclonal antibody against SARS-CoV-2 S protein (Sino Biological, 40150-D001) were plated at a final concentration of 1:5000. Samples were prepared in 2-fold serial dilutions starting at 1:4 and ending at 1:512. Blocking solution was removed, and diluted samples were added to the plates and incubated at room temperature for 2 hours. Plates were washed 3 times, and 50 µL of secondary antibody (1:5000 dilution of HRP-conjugated goat anti-human Fc-specific IgG; Invitrogen #A18823) was added to each well, and plates were incubated in the dark at room temperature for 1 hour. Plates were washed and all residual liquid was removed. SIGMAFAST OPD (o-phenylenediamine dihydrochloride) solution (Millipore Sigma) was added to each well, and plates were incubated in the dark for 10 minutes. 3M HCl (ThermoFisher) was added to each well to stop reaction, and the optical density of each plate was read at 490 nm using a SpectraMax i3 ELISA Plate Reader (BioTek Instruments). Cutoff values were calculated by adding the average of all negative control OD values and 3 times the standard deviation of the negative control values. Values were considered positive (responders) if at or above the cutoff value and negative (non-responders) if below the cutoff.

### Statistical analyses

Comparisons of clinical characteristics, infectious virus recovery, and between anti-S IgG responders and non-responders were tested using a two-sided Fisher’s exact test. Prior to conducting statistical analyses of anti-S IgG values, area under the curve (AUC) values were calculated by plotting the normalized optical density values against the sample dilution. A two-way ANOVA using Tukey’s multiple comparisons was used to assess differences in anti-S IgG AUC among groups, as well as differences in SARS-CoV-2 N Ct values among groups. Multivariate regression models (logistic and linear) were used to investigate the association of immunological measures (CPE, viral RNA level, and anti-Spike IgG) with pregnancy and vaccination, controlling for participant age, race/ethnicity, and area deprivation index (ADI) as necessary. An interaction term of the predictor variables was also included in the statistical models to allow for the predicted probabilities to vary by pregnancy and vaccination status. Contrasts of marginal effects were performed as post-estimation comparisons across pregnancy and vaccination groups. All analyses were performed using either Prism software version 9.5 (Graphpad) or using Stata version 17.0 (StataCorp).

## Results

### Clinical Data Analysis

Clinical outcomes between pregnant and non-pregnant women with confirmed SARS-CoV-2 infections differed. While pregnant women were less likely to report symptoms than non-pregnant women (OR = 0.41; CI = 0.23-0.71; *P* = 0.003); among symptomatic individuals, pregnant women were more likely to require hospitalization (OR = 4.2; CI = 2.0-8.6, *P* = 0.0003) or be admitted to the ICU (OR = 4.5; CI = 1.2-14.2, *P* = 0.02) with COVID-19 as their primary reason for admission (OR = 3.1; CI = 1.4-6.8; *P* = 0.009) (**Table 2**). In addition, pregnant women were more likely to be placed on supplemental oxygen therapy than non-pregnant women (OR = 3.1; CI = 1.3-6.9, *P* = 0.012) (**Table 2**).

**Table 2.** Differences in clinical severity between non-pregnant and pregnant females.

|  | **Pregnant** | *First Trimester* | *Second Trimester* | *Third Trimester* | **Non-Pregnant** | **Odds Pregnant vs. Non-Pregnant Total [CI]** | **P-value** |
| --- | --- | --- | --- | --- | --- | --- | --- |
| Variables, N | 117 | 28 | 36 | 53 | 335 |  |  |
| Symptomatic Total, n (%) | 91 (77.7%) | 25 (89.3%) | 32 (88.9%) | 34 (64.1%) | 300 (89.6%) | 0.41 [0.23-0.71] | 0.003 |
| Unvaccinated, n (%) | 63 (69.2%) | 18 (72.0%) | 24 (75.0%) | 21 (61.8%) | 220 (73.3%) | 0.33 [0.17-0.61] | 0.0014 |
| Vaccinated, n (%) | 28 (30.8%) | 7 (28.0%) | 8 (25.0%) | 13 (38.2%) | 80 (26.7%) | 0.77 [0.26-2.1] | 0.76 |
| Hospitalization Total, n (%) | 17 (14.5%) | 0 (0.0%) | 3 (8.3%) | 14 (26.4%) | 13 (3.9%) | 4.2 [2.0-8.6] | 0.0003 |
| COVID reason for admission Total, n (%) | 13 (11.1%) | 0 (0.0%) | 2 (5.6%) | 11 (20.7%) | 13 (3.9%) | 3.1 [1.4-6.8] | 0.009 |
| ICU Admittance Total, n (%) | 6 (5.1%) | 0 (0.0%) | 1 (2.8%) | 5 (9.4%) | 4 (1.2%) | 4.5 [1.2-14.2] | 0.02 |
| Supplemental O2, n (%) | 11 (9.4%) | 0 (0.0%) | 2 (5.6%) | 9 (17.0%) | 11 (3.3%) | 3.1 [1.3-6.9] | 0.012 |

### Distribution of SARS-CoV-2 variants among pregnant and non-pregnant women

Whole genome sequencing (WGS) results were used to classify infecting SARS-CoV-2 variants into one of five categories: ancestral lineages (i.e., those circulating prior to Alpha), Alpha variant, Delta variant, Omicron variant (through BA.2.12.1), and other (i.e., encompassing all other variants). Among unvaccinated individuals, most samples collected were from infections prior to vaccine availability and were predominately caused by ancestral lineages (40% in non-pregnant women and 32% in pregnant women); samples from infections by all other variants, however, were proportionally represented (**Table 3**). As emergency use authorization of both the Pfizer/BioNTech and Moderna mRNA-1273 vaccines coincided with the emergence and dominance of the Alpha variant, many samples collected from the vaccinated non-pregnant and pregnant cohort were individuals experiencing breakthrough infections from either the Delta variant (53% and 24%, respectively) or Omicron variants (38% and 73%, respectively) (**Table 3**).

**Table 3.** SARS-CoV-2 Lineage Distribution.

|  | **Ancestral** | **Alpha** | **Delta** | **Omicron** | **Other** | **Total** |
| --- | --- | --- | --- | --- | --- | --- |
| **Unvaccinated** |  |  |  |  |  |  |
| Non-pregnant, n (%) | 97 (40%) | 58 (24%) | 32 (13%) | 23 (9%) | 34 (14%) | 244 |
| Pregnant, n (%) | 27 (32%) | 12 (14%) | 14 (17%) | 23 (27%) | 8 (10%) | 84 |
| **Vaccinated** |  |  |  |  |  |  |
| Non-pregnant, n (%) | 5 (6%) | 1 (1%) | 48 (53%) | 35 (38%) | 2 (2%) | 91 |
| Pregnant, n (%) | 0 (%) | 0 (0%) | 8 (24%) | 24 (73%) | 1 (3%) | 33 |

### SARS-CoV-2 virus RNA level and recovery of infectious virus from upper respiratory samples

To evaluate if the differences in clinical severity between non-pregnant and pregnant women were due to differences in virus load, we compared infectious virus recovery and viral RNA levels (Ct values) for each group. Because there were no statistical differences in the days to symptom onset between symptomatic non-pregnant (2.2 ± 2.6 days) and pregnant (2.4 ± 3.4 days) women within this cohort, these analyses were conducted regardless of the days to symptom onset and whether the patient was symptomatic or asymptomatic at the time of collection, consistent with previous studies [17]. The number of samples from which infectious virus was recovered was significantly lower among non-pregnant vaccinated than unvaccinated women (*P*<0.05; **Figure 1A**). While a similar trend was noted between unvaccinated and vaccinated pregnant women, this did not reach statistical significance. There were no statistical differences in the rates of infectious virus recovery between non-pregnant and pregnant women, regardless of vaccination status. Viral RNA levels were similarly distributed between pregnant and non-pregnant women, and no statistical differences were observed (**Figure 1B**). Additionally, we assessed whether there were differences between the number of individuals with high (Ct > 20; low viral RNA levels) versus low (Ct ≤ 20; high viral RNA levels) viral RNA levels within each group. While greater percentages of vaccinated non-pregnant and pregnant women had lower viral levels (58% and 60%, respectively) than their unvaccinated counterparts (45% and 58%, respectively), these differences were not statistically significant (**Figure 1B, red text**).

**Figure 1.**
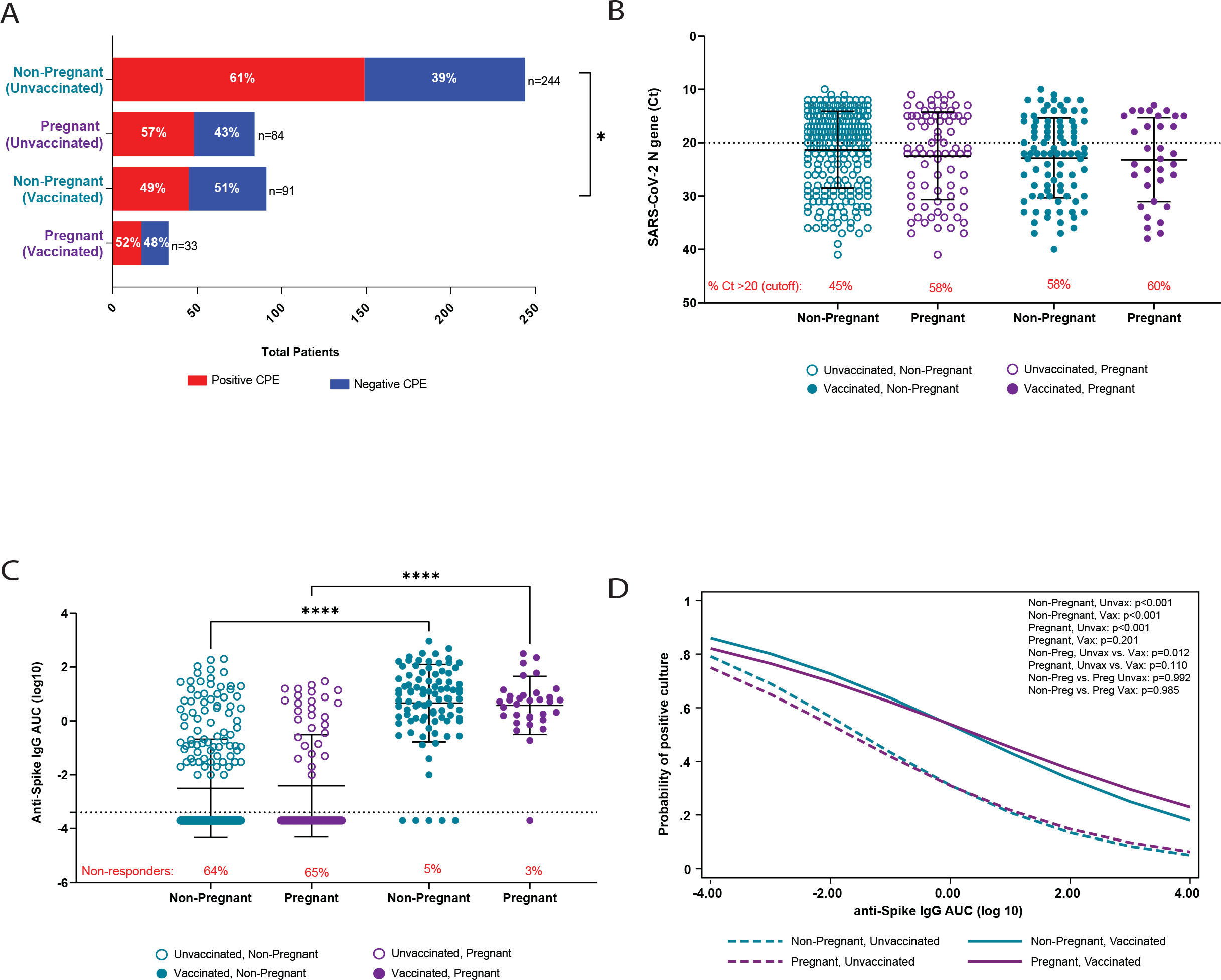
SARS-CoV-2 viral RNA levels and antibody responses stratified by pregnancy and vaccination status. Remnant clinical upper respiratory tract specimens were used determine rates of infectious virus recovery (A), viral RNA level (B), and anti-spike (ancestral spike) IgG titers (C) from mucosal swab samples. In (A), a positive cytopathic effect (CPE) in tissue culture was indicative of the presence of infectious virus. The *dashed line* in (B) represents the cutoff value (Ct ≤20) between high viral RNA and low viral levels, and the red text indicates the percentage of participants with Ct values > 20 (low viral RNA levels). The *dashed line* in (C) represents the limit of detection, and the red text indicates the percentage of non-responders (results below the limit of detection). Multivariate logistic regression was used to assess the correlation between anti-spike IgG titer and the probability of recovery of infectious virus (D). Analysis included Fisher’s exact test (A) and) and two-way ANOVAs with Tukey’s multiple comparisons test (B-C). \**P* < 0.05, *** *P* < 0.001, **** *P* < 0.0001. *anti-S IgG*, anti-ancestral strain spike immunoglobulin G; *AUC*, area under the curve; *Ct*, cycle threshold; *SARS-CoV-2*, severe acute respiratory syndrome coronavirus 2.

### Comparisons of mucosal anti-S IgG titers between pregnant and non-pregnant women

Although previous reports suggest that pregnant women have reduced antibody responses to SARS-CoV-2 infection [6, 28-30], these studies focused solely on serum antibody responses. As SARS-CoV-2 infection initiates in the upper respiratory tract, we sought to evaluate whether differences in mucosal IgG responses between non-pregnant and pregnant women may account for differences in clinical severity. Vaccinated individuals had greater anti-S IgG titers than unvaccinated individuals, regardless of pregnancy status (*P* < 0.0001; **Figure 1C**). Proportions of individuals with undetectable anti-S IgG (i.e., non-responders) were greater in unvaccinated women compared to vaccinated women (non-pregnant: *P* < 0.0001; pregnant: *P* < 0.0001), but there were no statistically significant differences between pregnant and non-pregnant women within vaccination groups (**Figure 1C, red text**). The correlation between anti-S IgG titers and infectious virus recovery and between anti-S IgG titers and viral RNA Ct values was examined as a proxy to assess whether there were differences in the antiviral activity of antibodies produced by non-pregnant and pregnant women. In the regression model controlling for age, race/ethnicity, and ADI, there was a strong inverse correlation between anti-S IgG AUC and the probability of recovering infectious virus (**Figure 1D**) as well as viral RNA level (**Supplemental Figure 1A**) among unvaccinated women, regardless of pregnancy status, and among vaccinated non-pregnant women. While similar inverse relationships were observed for vaccinated pregnant women, they were not statistically significant. Notably, when the variable for time post-symptom onset was included in the regression models (excluding asymptomatic individuals; non-pregnant, N=35; pregnant, N=26), the inverse correlation between anti-S IgG AUC and the probability of recovering infectious virus **(Supplemental Figure 1B)** as well as between anti-S IgG and viral RNA Ct values (not shown) remained unchanged.

### Age and trimester of pregnancy influence mucosal immunity in pregnant, vaccinated women

To further interrogate possible pregnancy-associated differences in mucosal antibody responses and viral level, we determined whether maternal age or gestational age contributed to observed variability in SARS-CoV-2 anti-spike IgG AUC values (**Figure 1C**). Individuals in our cohort were classified into one of three maternal age groups: ages 18-24, ages 25-34, and ages 35-44. We first compared viral RNA level, mucosal anti-S IgG AUC values, and rate of infectious virus recovery regardless of days to symptom onset or whether the patients were symptomatic or asymptomatic. Among unvaccinated individuals, viral RNA level (**Figure 2A**), rates of infectious virus recovery (**Figure 2A, red text**), and mucosal anti-IgG AUC values (**Figure 2B**), were similar across all groups, regardless of pregnancy status. No age-related differences were noted in either viral RNA level **(Figure 2A**) or anti-S IgG AUC values (**Figure 2B**) among non-pregnant, vaccinated women. Among vaccinated, pregnant women, viral RNA levels increased (**Figure 2A**) and anti-S IgG AUC values decreased (**Figure 2B**) with maternal age, with pregnant women ages 25-34 and ages 35-44 having significantly greater viral RNA (*P* < 0.001) and lower anti-S IgG AUC values (*P* < 0.001) compared to pregnant women ages 18-24 (**Figure 2A-B**). Additionally, we observed trends in which the rates of infectious virus recovery decreased with age in non-pregnant, vaccinated women, but increased with age in pregnant, vaccinated women although these trends did not reach statistical significance (**Figure 2A, red text**). The proportion of non-responders was greater in unvaccinated women compared to vaccinated women, regardless of age (non-pregnant: *P* < 0.0001; pregnant: *P* < 0.0001), but there were no statistically significant age-associated differences between pregnant and non-pregnant women within vaccination groups (**Figure 2B, red text**). When controlled for race/ethnicity and ADI, similar trends in the relationship between Ct values and age (**Supplemental Figure 2A**) and with the probability of recovery of infectious virus and age (**Supplemental Figure 2B**) within each group were observed, however, none were statistically significant. Likewise, when controlled for race/ethnicity, ADI, and time since vaccination (**Supplemental Figure 1C**), we observed a similar age-associated trend of decreased anti-S IgG AUC values among vaccinated pregnant women which was not statistically significant. Importantly, the average time between completion of vaccination and infection was similar among non-pregnant (176 ± 85 days) and pregnant (187 ± 95 days) women.

**Figure 2.**
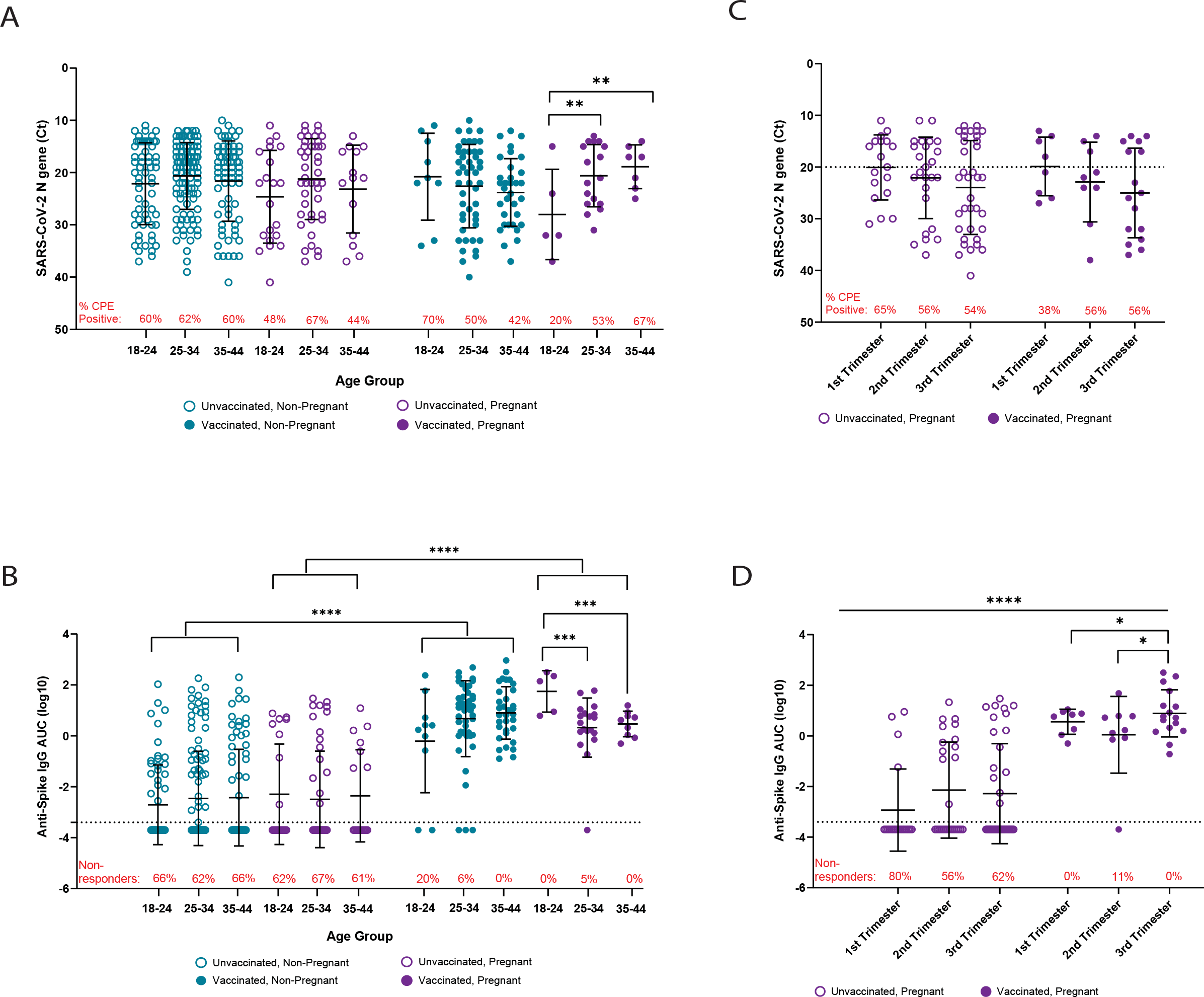
The effects of maternal and gestational age on mucosal viral RNA levels and antibody responses. (A-B) Study participants were divided into three maternal age groups: ages 18-24, ages 25-34, and ages 35-44, and samples were re-analyzed to assess differences in viral RNA levels (A) and anti-S IgG (B). (C-D) Results from unvaccinated and vaccinated pregnant women were stratified according to trimester of pregnancy and re-analyzed to assess differences in viral RNA levels (C) and anti-S IgG (D). The red text in (A,C) indicates the percentage of individuals with recoverable infectious virus, and the red text in (B,D) indicates the percentage of non-responders (i.e. those with anti-S IgG below the limit of detection). (A-D): Two-way ANOVA with Tukey’s multiple comparisons test. \**P* < 0.05, \*\**P* < 0.01, *\*\*\*P* < 0.001, *\*\*\*\*P* < 0.0001.

Next, we examined the relationships between gestational age, viral RNA level, mucosal anti-S IgG AUC values, and recovery of infectious virus, regardless of days to symptom onset or whether the patients were symptomatic or asymptomatic. Although no statistical differences in viral RNA level (**Figure 2C)** or recovery of infectious virus (**Figure 2C, red text)** were observed across trimesters of pregnancy, a trend of reduced viral level across trimester was observed, with the lowest values being recorded in the third trimester for both unvaccinated and vaccinated pregnant women. Among vaccinated pregnant women, anti-S IgG AUC values were greater in the third trimester compared to either the first (*P* < 0.05) or second (*P* < 0.05) trimester of pregnancy (**Figure 2D**). Proportions of non-responders (i.e., those with undetectable anti-S IgG) within each trimester were greater in unvaccinated compared to vaccinated pregnant women (1^st^ trimester: *P*=0.0002; 2^nd^ trimester: *P*=0.02; 3^rd^ trimester: *P*=0.002); and were not statistically different between trimesters within vaccination groups (**Figure 2D, red text**). When controlled for race/ethnicity, ADI, and days post-symptom onset for symptomatic individuals (**Supplemental Figure 2C**), a similar trimester-associated decrease in viral RNA level was observed, but this did not reach statistical significance. When controlled for race/ethnicity, ADI, and time between completion of vaccination and infection (**Supplemental Figure 1D**), a trimester-associated increase in anti-S IgG was observed, but it was not statistically significant. The mean time between completion of vaccination and infection was similar between women in their first trimester (216 ± 57 days) and second trimester (214 ± 105 days) but decreased in women in their third trimester (159 ± 102 days) of pregnancy. Taken together, these results suggest that mucosal antibody responses to SARS-CoV-2 infection are reduced with increased maternal age and earlier in pregnancy.

### Pregnant women infected with Omicron variants have reduced mucosal anti-S IgG levels

This patient cohort included individuals infected with both Delta and Omicron (through BA.2.12.1) variants. We conducted an additional analysis of pregnancy-associated differences based on the infecting variant. No differences in viral RNA level were detected among either pregnant or non-pregnant women (**Figure 3A**). Pregnant, vaccinated individuals infected with Omicron, but not Delta, variants had significantly lower anti-S IgG AUC values than non-pregnant, vaccinated women (*P* < 0.05; **Figure 3B**). In contrast, anti-S IgG AUC values were comparable between unvaccinated pregnant and non-pregnant women infected with either Delta or Omicron variants. The proportion (**Figure 3B, red text**) of unvaccinated, non-pregnant women with non-detectable anti-S IgG titers was lower among those infected with Omicron variants compared to Delta (*P* = 0.01) but was higher among unvaccinated pregnant women (*P* = 0.0003). Similar observations were made among vaccinated individuals but were not statistically significant.

**Figure 3.**
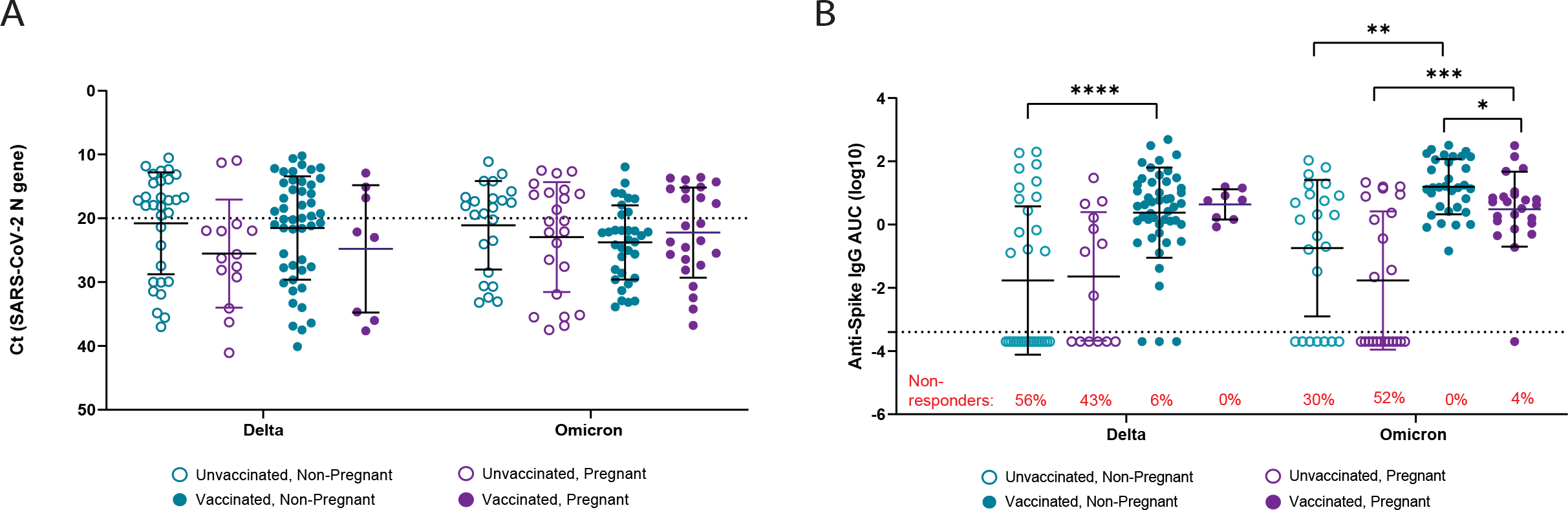
Analysis of mucosal viral RNA levels and antibody responses to Delta and Omicron breakthrough infections during pregnancy. Samples were classified according to infecting strain (delta or omicron), pregnancy status, and vaccination status, and re-analyzed to assess differences in viral RNA level (A) and anti-S (ancestral/vaccine strain) IgG (B). Two-way ANOVA with Tukey’s multiple comparisons tests (A-B) \**P* < 0.05, \*\**P* <0.01, *\*\*\*P* < 0.001, *\*\*\*\*P* <0.0001.

In our previous studies, we have shown that lower Ct values (Ct≤20; i.e., high viral RNA) are associated with greater rates of infectious virus recovery [14, 17], and that individuals infected with Delta variants had greater rates of infectious virus recovery than those infected with Omicron variants [17]. Thus, we assessed whether there were differences in infectious virus recovery among individuals with high viral RNA levels, but found no statistical differences in the rates of infectious virus recovery between individuals infected with either the Delta or Omicron variants within this cohort (**Supplemental Figure 2D**).

## Discussion

The ongoing COVID-19 pandemic has raised awareness about pregnant women being at greater risk for severe complications arising from many viruses, including SARS-CoV-2 [2, 5, 31]. In a retrospective cohort of pregnant and non-pregnant women with confirmed SARS-CoV-2 infection, we observed that disease severity, including ICU admission and oxygen supplementation, was greater among pregnant than non-pregnant women. We further explored the role of vaccination in mucosal immunity and recovery of live SARS-CoV-2 and viral RNA from the upper respiratory tract. Vaccination reduced recovery of infectious virus in non-pregnant, but not pregnant, women suggesting that vaccine-induced immunity and protection might be reduced during pregnancy, as previously reported for other infectious diseases [2]. Greater maternal age was associated with reduced mucosal antibody responses and greater viral RNA levels, especially among pregnant women infected with the Omicron variant. These findings provide mechanistic insights into how pregnant women are at greater risk of severe COVID-19, including from breakthrough infections with variants of concern following receipt of the monovalent COVID-19 vaccines.

Advanced maternal age (i.e., 35+ years of age) has previously been associated with severe clinical outcomes and adverse fetal outcomes following SARS-CoV-2 in pregnant women [32, 33]. While several studies highlight that SARS-CoV-2 infected pregnant women typically present with asymptomatic or mild infections [3, 34], there are data illustrating that pregnant women with SARS-CoV-2 infections are at increased risk of hospitalization, ICU admittance, invasive ventilation, and death than non-pregnant women [3, 5, 31, 35]. Increased risk of severe outcomes among pregnant women have persisted at least through the Delta variant wave [5], and we now show that this further persists in the Omicron variant wave of the pandemic.

Existing serological evidence in SARS-CoV-2 infection demonstrates that pregnant women have enhanced inflammatory responses and reduced humoral responses compared to non-pregnant women [6, 8, 36, 37]. The data from the current study add to the existing literature by showing that mucosal antibody responses also are reduced in pregnant women compared with non-pregnant women, with further reductions with advanced maternal age, in the first trimester of pregnancy, and with some infecting variants of concern.

### Clinical Implications

Among pregnant women with confirmed SARS-CoV-2 infection, reduced mucosal antibody responses were associated with greater infectious virus recovery and viral RNA levels, especially among women with advanced maternal age and women infected with the Omicron variant. These data highlight that monovalent vaccines were not sufficient to protect pregnant women against Omicron, which is consistent with reports in the general population [38-41], and highlight the need for receipt of the bivalent booster in pregnant women. Pregnant women were not included in phase III clinical trials for any of the vaccine candidates or the bivalent booster [42]; and limited data are available from women who became pregnant while participating in vaccine trials [43-46]. Because pregnancy is a unique biological state [47-51], additional studies evaluating vaccine efficacy and the use of SARS-CoV-2 therapeutic agents (including use of monoclonal antibodies) are necessary to ensure that the same correlates of protection apply to this high-risk population [52]. Currently, the CDC recommends that pregnant women consult with their physician to make decisions on vaccination. However, the lack of supporting vaccine safety and efficacy in pregnancy complicates the benefit-risk analysis for healthcare providers and pregnant women. Greater use of animal models to assess vaccine efficacy during pregnancy and how pregnancy may alter vaccine-induced immunity and protection from breakthrough infection is needed [42].

### Study Limitations

The primary limitation of this study is the small sample size. While these studies were powered for the primary clinical outcome, we were unable to give adequate statistical consideration for additional potential confounding variables (e.g., time since symptom onset, time between vaccination and sample collection) in the regression models. This was due both to incomplete charting data (e.g. 77 symptomatic participants without a reported date of symptom onset), and due to the use of convenience samples which limited our ability to control for race/ethnicity, age, ADI, and time between vaccination and sample collection. When we applied multivariate regression analysis that controlled for these variables, the trends in our data remained consistent but lost statistical power. This highlights the need to verify these data in a larger clinical cohort. Moreover, only upper respiratory samples were collected, and no serum samples were available for additional analyses (e.g., IgA antibody levels, virus neutralization or cross-reactivity with Spike proteins from variants of concern). For clinical outcomes, pregnant women in our study were reportedly less symptomatic than non-pregnant women; this was, however, based on self-reporting from a general list of questions that may not distinguish COVID-19-related illness from pregnancy-associated symptoms (e.g., fatigue, muscles or body aches, headache, digestive issues, nausea, or vomiting). Symptomatic COVID-19 cases among pregnant women may not be accurately represented. Because samples were collected at different points of care within the Johns Hopkins Medical System, differences in sample collection may contribute to the variability in infectious virus recovery, viral RNA levels, and antibody titers.

## Conclusions

Pregnancy is associated with more severe outcomes from COVID-19 during the Omicron wave of the pandemic. Advanced maternal age, first trimester of pregnancy, and infection with Omicron were identified as factors contributing to decreased mucosal antibody responses with concomitant increases in live virus recovery and mucosal viral RNA levels. Greater consideration of pregnancy in prophylactic and therapeutic interventions for people infected with SARS-CoV-2 [53] is needed to enable pregnant women and their healthcare providers to make evidence-based decisions about care.

## Data Availability

All data produced in the present study are available upon reasonable request to the authors.

## Author Contributions

*Concept and Design*: LAS, REE, JS, ALC, IB, SLK, EYK, AP, HHM

*Acquisition, analysis, and interpretation of data*: LAS, REE, JS, AY, AF, CPM, JMN, MF, OA, SD, CB, AP, HHM, EYK, SLK

*Drafting of Manuscript*: LAS, AY, SLK

*Critical revision of the manuscript for important intellectual content*: All authors.

*Statistical analysis*: LAS, AY, REE, JS

*Obtained Funding*: SLK, ALC, AP, HHM

*Supervision*: SLK, EYK, AP, HHM

## Acknowledgements

We would like to sincerely thank Barbara Wesley, MD, MPH from the Center for Drug Evaluation and Research at the U.S. Food and Drug Administration for her thoughtful suggestions during conceptualization and design and during preparation of this manuscript.

## Figure Legends

**Supplemental Figure 1 – Multivariate analysis of SARS-CoV-2 antibody responses**.

Multivariate logistic regression was used to assess the correlation between anti-spike IgG titer and viral RNA levels (A), anti-spike IgG titer and the probability of infectious virus recovery (B), anti-spike IgG titer and age (C), and trimester of pregnancy and anti-spike IgG titer (D). (A-C): Variables were continuous, and p-values represent strength of correlation between variables for each categorical group and comparisons between groups. (D): Trimester of pregnancy was utilized as a categorical variable, p-values represent comparisons between stated groups.

**Supplemental Figure 2 – Multivariate analysis of SARS-CoV-2 viral RNA levels and recovery of infectious virus**.

Multivariate logistic regression was used to assess the correlation between age and viral RNA levels (A), age and the probability of infectious virus recovery (B), viral RNA levels and trimester of pregnancy (C). Recovery of infectious virus from samples with high viral RNA levels (Ct ≤ 20) is reported in (D). (A-B) Variables were continuous, and p-values represent strength of correlation between variables for each categorical group and comparisons between groups. (C): Trimester of pregnancy was utilized as a categorical variable, p-values represent comparisons between stated groups. (D): A Fisher’s exact test was utilized.

