## Supplementary figures and images for "Reduced control of SARS-CoV-2 infection is associated with lower mucosal antibody responses in pregnant women"

### Supplemental Figures

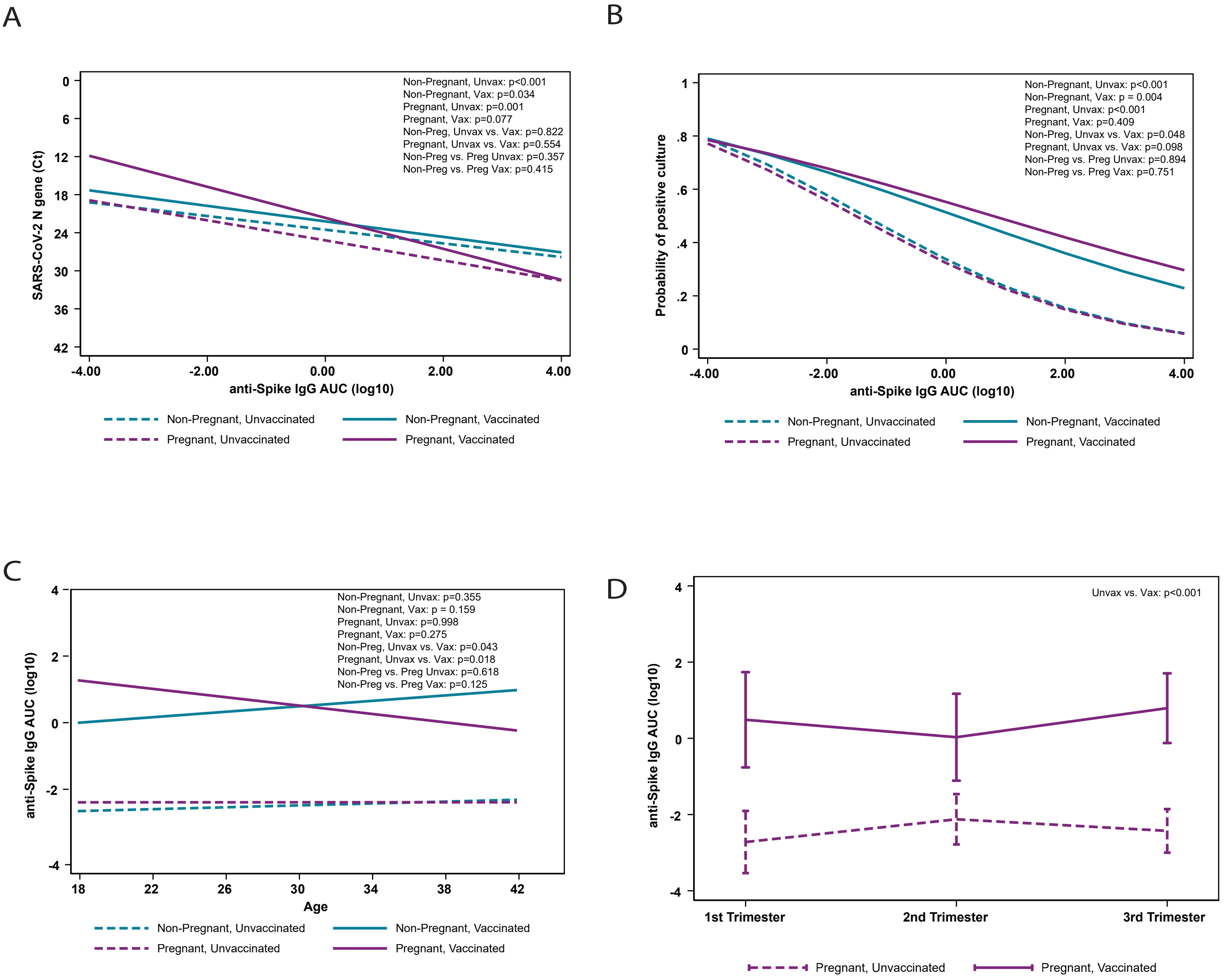


St Clair et al., Supp Fig 1

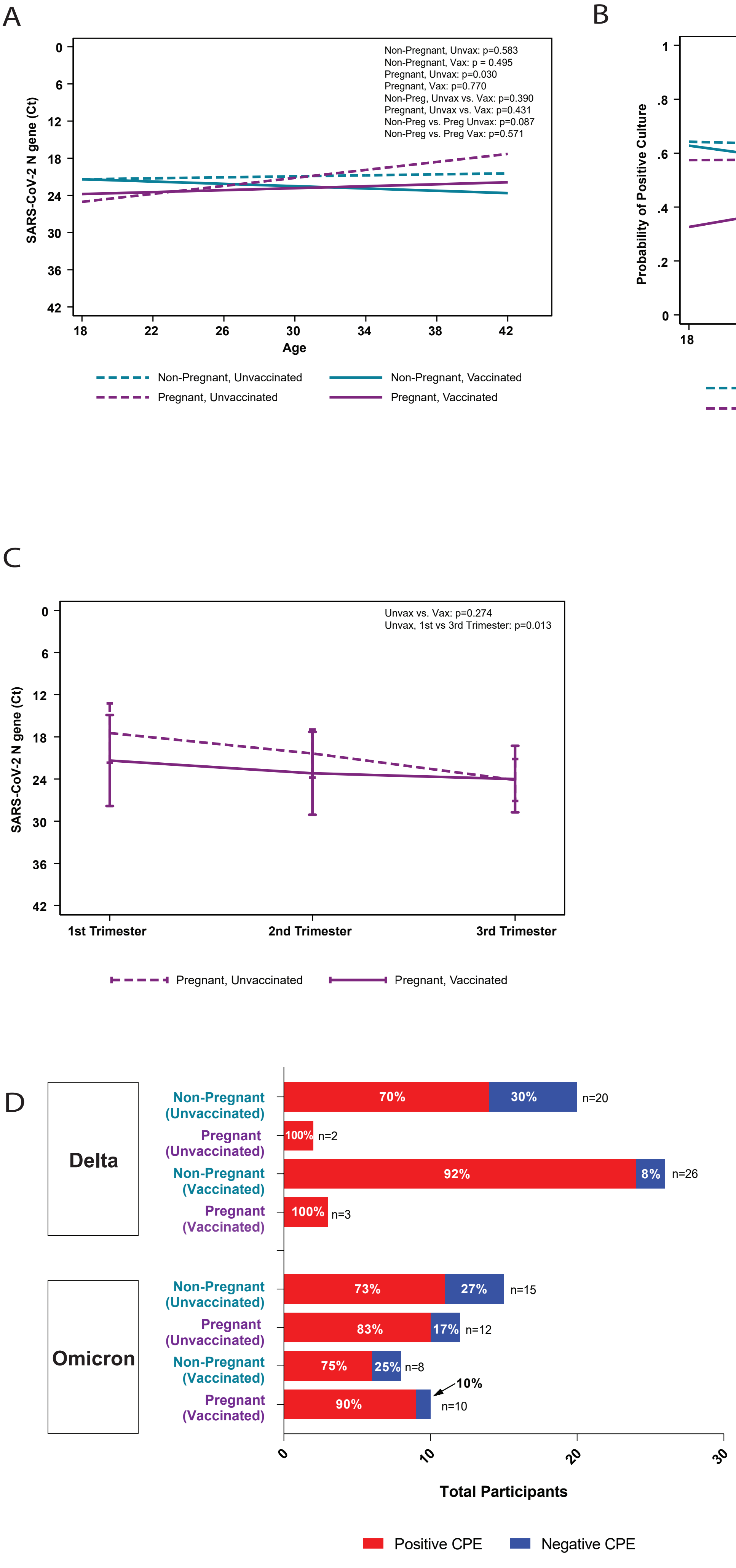


St Clair et al., Supp Fig 2
